# Association study of human leukocyte antigen (HLA) variants and idiopathic pulmonary fibrosis

**DOI:** 10.1101/2023.07.20.23292940

**Authors:** Beatriz Guillen-Guio, Megan L. Paynton, Richard J. Allen, Daniel P.W. Chin, Lauren J. Donoghue, Amy Stockwell, Olivia C. Leavy, Tamara Hernandez-Beeftink, Carl Reynolds, Paul Cullinan, Fernando Martinez, CleanUP-IPF Investigators of the Pulmonary Trials Cooperative, Helen L. Booth, William A. Fahy, Ian P. Hall, Simon P. Hart, Mike R. Hill, Nik Hirani, Richard B. Hubbard, Robin J. McAnulty, Ann B. Millar, Vidya Navaratnam, Eunice Oballa, Helen Parfrey, Gauri Saini, Ian Sayers, Martin D. Tobin, Moira K. B. Whyte, Ayodeji Adegunsoye, Naftali Kaminski, Shwu-Fan Ma, Mary E. Strek, Yingze Zhang, Tasha E. Fingerlin, Maria Molina-Molina, Margaret Neighbors, X. Rebecca Sheng, Justin M. Oldham, Toby M. Maher, Philip L. Molyneaux, Carlos Flores, Imre Noth, David A. Schwartz, Brian L. Yaspan, R. Gisli Jenkins, Louise V. Wain, Edward J. Hollox

## Abstract

**Introduction:** Idiopathic pulmonary fibrosis (IPF) is a chronic interstitial pneumonia marked by progressive lung fibrosis and a poor prognosis. Recent studies have highlighted the potential role of infection in the pathogenesis of IPF and a prior association of the *HLA-DQB1* gene with idiopathic fibrotic interstitial pneumonia (including IPF) has been reported. Due to the important role that the Human Leukocyte Antigen (HLA) region plays in the immune response, here we evaluated if HLA genetic variation was associated specifically with IPF risk.

**Methods:** We performed a meta-analysis of associations of the HLA region with IPF risk in individuals of European ancestry from seven independent case-control studies of IPF (comprising a total of 5,159 cases and 27,459 controls, including the prior study of fibrotic interstitial pneumonia). Single nucleotide polymorphisms, classical HLA alleles and amino acids were analysed and signals meeting a region-wide association threshold *p*<4.5×10^−4^ and a posterior probability of replication >90% were considered significant. We sought to replicate the previously reported *HLA-DQB1* association in the subset of studies independent of the original report.

**Results:** The meta-analysis of all seven studies identified four significant independent single nucleotide polymorphisms associated with IPF risk. However, none met the posterior probability for replication criterion. The *HLA-DQB1* association was not replicated in the independent IPF studies.

**Conclusion:** Variation in the HLA region was not consistently associated with risk in studies of IPF. However, this does not preclude the possibility that other genomic regions linked to the immune response may be involved in the aetiology of IPF.

## INTRODUCTION

Idiopathic pulmonary fibrosis (IPF) is a lung disease characterised by progressive scarring of the alveoli leading to impaired gas exchange. The median survival after IPF diagnosis is 2-5 years and there are limited drug treatments for patients (1). Several studies have reported both environmental and genetic risk factors related to IPF (2). However, the cause of IPF development remains unclear.

Genetic variation in the Human Leukocyte Antigen (HLA) region, also known as the major histocompatibility complex (MHC) region, has been reported as associated with inflammatory and respiratory diseases (3,4). This includes fibrotic idiopathic interstitial pneumonia (fIIP), where *HLA-DQB1*06:02* has been associated with increased risk of disease (5). Additionally, *HLA-DRB1*15:01* has been found to be more prevalent among IPF patients than controls (6). The mechanism behind these associations is unclear. However, it has been previously suggested that respiratory infections, including coronavirus infectious disease 2019 (COVID-19), could trigger the development and progression of interstitial lung diseases, including IPF (7-9). Indeed, antiviral drugs against herpesviruses have been proposed to attenuate disease progression in IPF patients (10). Given the important role of the MHC receptors encoded by HLA genes in presenting viral antigens to the host immune system, genetic variation at the HLA region could influence the response to these infections and therefore IPF pathophysiology.

Recent studies on large biobanks have emphasised the pleiotropy of the HLA region (11). This region is highly polymorphic and gene-dense, making the interpretation of single nucleotide polymorphisms (SNP) associations difficult, and its study requires specific imputation techniques (12). In this sense, the latest SNP imputation panels enable improved imputation accuracy within the HLA region. Additionally, specific HLA imputation panels also allow the imputation of classical HLA alleles and amino acid alleles across the HLA region, which are biologically informative (13).

Given the potential role of infection response in IPF physiopathology, and the suggested association of *HLA-DQB1*06:02* with disease, here we present an analysis of the HLA region across seven independent case-control cohorts for IPF, with a total of over 5,000 cases and 27,000 controls. We used an HLA-specific imputation approach to impute SNP variation, classical HLA alleles and amino acid alleles (hereafter to be known as variants) with the aim of identifying novel IPF risk loci within the HLA region.

## METHODS

### Sample, genotyping and quality controls

We analysed genomic data from seven previously described independent case-control studies, named here as CleanUP-UCD [Study of Clinical Efficacy of Antimicrobial Therapy Strategy Using Pragmatic Design -University of California Davis] (14,15), Colorado (16), Genentech (17,18), IPF-JES [Job Exposures Study] (19), UK (20), US (21) and UUS [United States, United Kingdom, and Spain] (22). CleanUP-UCD, IPF-JES, UK, US and UUS studies included patients diagnosed with IPF as cases and population controls. The Colorado study used cases of fIIP (including IPF cases among other conditions) and population controls and was the study that previously reported the fIIP association for *HLA-DQB1*06:02* (5). The Genentech study included patients with IPF and controls from non-IPF clinical trials of age-related macular degeneration, diabetic macular oedema, multiple sclerosis, asthma, and inflammatory bowel disease. In all seven studies, cases were diagnosed according to the American Thoracic Society and European Respiratory Society guidelines (23,24). The studies were performed in accordance with The Code of Ethics of the World Medical Association (Declaration of Helsinki) and approved by the appropriate institutional review or Research Ethics Committee. Further details about the seven studies can be found in the Supplement.

In Colorado, IPF-JES, UK, US, UUS, and CleanUP-UCD studies, individuals were genotyped using SNP arrays (Affymetrix and Illumina Inc.). Genotyping and quality control procedures have been previously detailed (20,22,25). In summary for the six studies, individuals were filtered based on low call rate estimates, sex mismatches, excess heterozygosity, relatedness, and non-European ancestry (based on principal components [PC] clustering with European individuals from The 1000 Genomes project). Individuals overlapping between studies were removed. Genotypes from individuals in the Genentech study were obtained from whole-genome sequencing using HiSeq X Ten platform (Illumina Inc.) to an average read depth of 30X. Related individuals and those with genotype missingness >10% or excess heterozygosity were excluded. Ancestry was determined using ADMIXTURE v1.23 (26), and individuals with a European genetic ancestry score > 0.7 were included in the analyses.

### Variant imputation in the HLA region and association tests

For each study, classical HLA alleles, amino acid alleles (AA) and SNPs located within chr6:28,510,120-33,480,577 (GRCh38) were imputed to capture genetic variation in the HLA region.

For Chicago, Colorado, UK, UUS, CleanUP-UCD, and IPF-JES studies, phasing and SNP Imputation to the TOPMed reference panel was performed using the TOPMed Imputation Server (6). Classical HLA alleles from class I genes (-A, -B, C) and class II genes (-DPA, -DPB, -DQA, -DQB, -DRB), amino acid (AA) alleles, and additional SNPs were imputed to the type one diabetes genomics consortium (T1DGC) panel using IMPUTE2 (V2.3.2) after chromosome phasing with SHAPE-IT v2.837 (27). After SNP imputation using both TOPMed and T1DGC panels, duplicated SNPs were removed, prioritising those imputed with TOPMed. For the Genentech study, variants with a minor allele frequency (MAF)<1% or ambiguous and SNPs absent from the TOPMed reference panel were removed. Classical HLA alleles and amino acid alleles were imputed using HLA-HD (28), and data was analysed using MiDAS (29).

Logistic regression analyses were performed in each study separately assuming an additive model. For Chicago, Colorado, UK, UUS, CleanUP-UCD, and IPF-JES studies, models were adjusted for the ten leading PCs to correct for population stratification. For the Genentech study, sex, age, and five genetic-ancestry PCs were included as covariates and reverse regression (30) was applied to flag any associations driven by the differential allele frequency of a control indication(s) rather than IPF. In all studies, low frequency variants (MAF < 1%) and variants with a poor imputation quality (r^2^ < 0.3) were removed from the analyses. A fixed-effect weighted meta-analysis combining the association results of the seven studies was performed using METAL (31) to establish the genetic variants associated with IPF. Variants were required to be present in at least two studies to be included in the analysis. The significance threshold was declared at *p*=4.50×10^−4^ after Bonferroni correction based on the number of independent SNPs, amino acid changes, and classical HLA alleles in this region. The number of independent signals in the region was identified using LD-prune from PLINK v1.9 (r^2^=0.2) (32). Conditional analyses were performed with COJO-GCTA (33) to identify independent sentinel variants. We used the Meta-Analysis Model-Based Assessment of Replicability (MAMBA) to assess the consistency of association results across studies (34). This tool calculates a posterior probability of replicability (PPR) that a given SNP has a non-zero replicable effect, indicating the likelihood of a specific SNP to replicate. Associations with a PPR >90% were considered consistent and likely to replicate. The Manhattan and conditional analyses plots were obtained using HLA-TAPAS (35) and R v4.0.0, respectively. Forest plots were performed with the *forestplot* R v4.1.3 package. Replication of the previously association with *HLA-DQB1*06:02* was sought using the 6 independent datasets (i.e., excluding Colorado study). Sensitivity analyses of all significant signals were conducted excluding the Colorado study, given that the case definition used included other fIIPs in addition to IPF.

## RESULTS

A total of 32,618 unrelated individuals of European ancestry (5,159 cases and 27,459 controls, Table S1) and 44,713 common variants within the HLA region (43,544 SNPs, 174 classical alleles, and 995 AA) were included in the analyses. After meta-analysis, four independent loci met the HLA-wide significance threshold (*p*=4.50×10^−4^) (Table 1, Table S2, Figures 1 and 2).

**Table 1:**
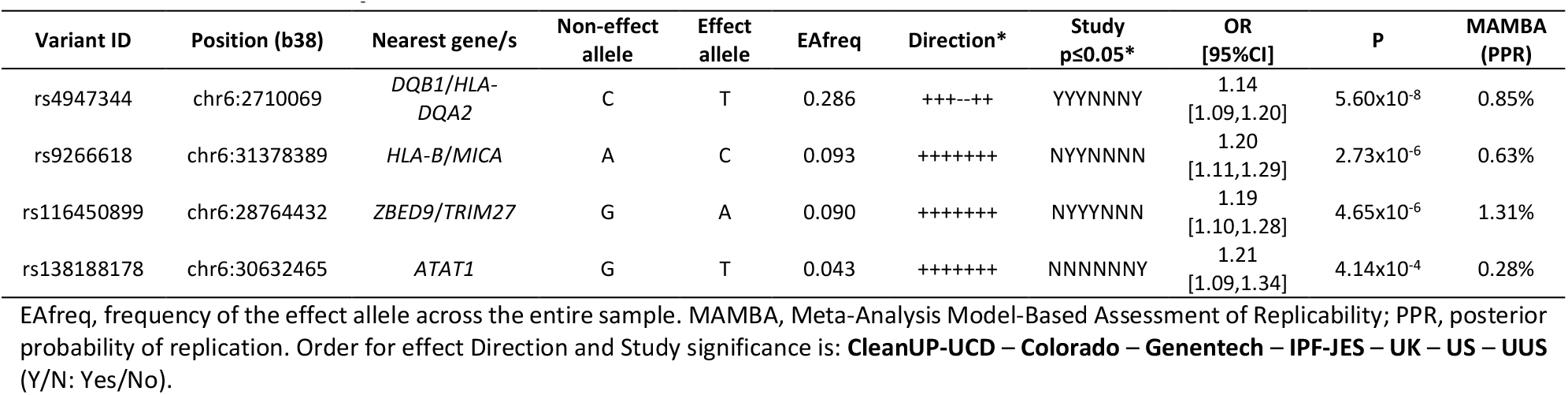
Association analysis results for sentinel variants.

**Figure 1.**
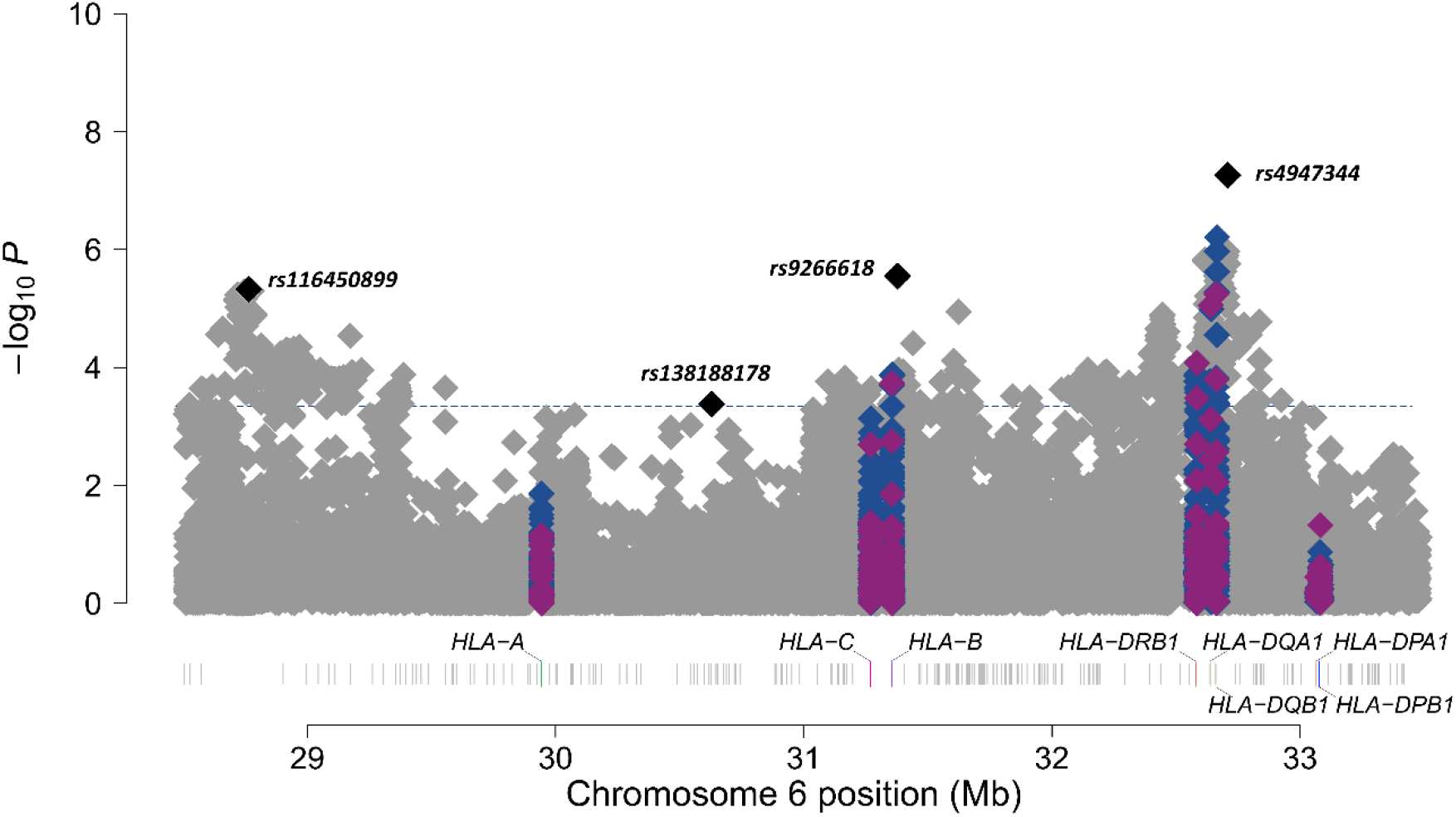
Manhattan plot of meta-analysis results at chromosome 6 region. SNPs are shown in grey, amino acids in blue, and classical HLA alleles in purple. The four sentinel variants are highlighted in black. The y-axis shows the transformed p-values (–log10) while the x-axis represents chromosome positions in Mb (GRCh38/hg38). The horizontal line corresponds to the significance threshold of the study after Bonferroni correction (*p*=4.50×10^−4^).

**Figure 2.**
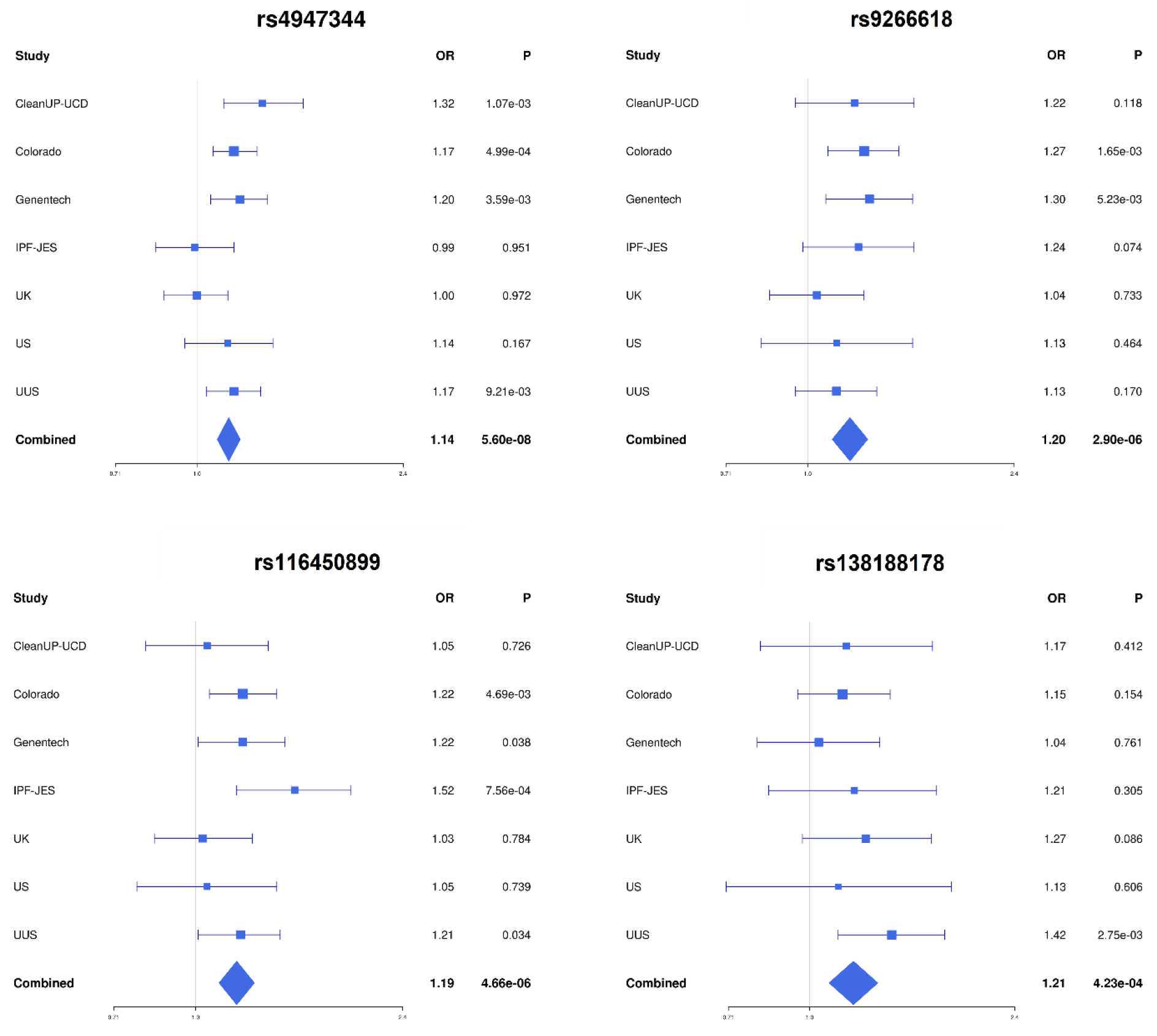
Forest plots of the association analysis results for the four sentinel variants.

The top sentinel variant was the SNP rs4947344, an intergenic variant located between *HLA-DQB1* and *HLA-DQA2* that was associated with IPF risk in our study (odds ratio (OR) = 1.14, 95% confidence interval (CI) = 1.09-1.20, *p*=5.60×10^−8^). After conditional analyses, we found that this variant was in linkage disequilibrium with the allele *HLA-DQB1*06:02*, previously associated with fIIP (5) (Figure S1).

The 3 remaining sentinel variants identified in the meta-analysis results were SNPs rs9266618, rs116450899, and rs138188178 (Table 1, Figures 1 and 2). rs9266618 and rs116450899 are intergenic variants located between *HLA-B* and the MHC Class I Polypeptide-Related Sequence A (*MICA*), and between the zinc finger BED-type containing 9 (*ZBED9)* and the tripartite motif containing 27 (*TRIM27*) genes, respectively, while rs138188178 is intronic to the gene encoding the Alpha Tubulin Acetyltransferase 1 (*ATAT1*).

None of the variants met nominal significance (*p*<0.05) in all seven contributing studies and the replicability assessment with MAMBA revealed that none of these association signals were considered likely to replicate (PPR<90% in all cases) (Table 1, Figure 2).

After sensitivity analyses excluding the Colorado study, three signal associations reached the Bonferroni P threshold, although the likelihood of replication remained very small for all three (Table S3). The association of *HLA-DQB1*06:02* with IPF was not independently replicated in the six independent IPF datasets (*p*=0.043, PPR= 0.09%) (Table S4, Figure S2).

## DISCUSSION

In this study, we used genotype data and imputation of SNPs, amino acids and classical HLA alleles from seven independent case-control cohorts to test for association with IPF susceptibility. Although we identified four significant associations, including a signal correlated with a previously reported association for *HLA-DQB1*06:02*, the effects were inconsistent across the contributing studies and this heterogeneity was reflected in a poor posterior probability for replication for all signals. The reason for this heterogeneity is unclear, but could be due to a broader definition of cases (including non-IPF fIIP) in the Colorado study (5) or use of a different methodology (whole genome sequencing) in the Genentech study, among other possibilities. While control subjects in the Genentech study were selected from clinical trials of non-IPF diseases, including those strongly associated with variation in the HLA region (e.g., asthma), reverse regression on these four associations did not detect that the control cohort composition was driving the IPF effect estimates (18).

We acknowledge several limitations. First, our study was restricted to individuals of European genetic ancestries. Thus, we cannot exclude the possibility that HLA variation might contribute to IPF susceptibility in individuals of other genetic ancestries. Next, the heterogeneity of signals across cohorts might suggest that HLA variation could contribute to forms of pulmonary fibrosis that do not meet the strict diagnostic criteria for IPF. Due to technology limitations, we were unable to evaluate all types of classical HLA alleles, which could have concealed potential IPF associations. Finally, the HLA is a complex region with known genetic and environmental interactions. Therefore, our approach of testing individual HLA variants with disease may be too simplistic to capture the complex interplay of these proteins with disease risk.

Our meta-analysis approach has the advantage of increasing the sample size for genetic studies of IPF. We restricted our analysis to studies of clinically defined IPF to avoid the issues of imprecise electronic healthcare record coding and resulting effect size attenuation that has been previously reported (36,37). By combining all available datasets, we were able to improve power to detect associations of large and modest effect sizes but at the expense of reserving samples for independent replication. To mitigate this, we used an approach that considers heterogeneity of association results across the meta-analysis to provide a posterior probability of replication. This approach allowed us to provide a quantitative indicator of the combined strength of the associations across studies rather than relying on a single combined P value which can be influenced by effects in individual studies.

Our results strongly suggest that individual genetic variants within the HLA are not associated with susceptibility to IPF in individuals with European ancestries, although they may have a role in other IIPs. Future studies investigating the role of infection in the aetiology of IPF should aim to assess the relationship of interactions of immune system genes.

## Data Availability

All data produced in the present study are available upon reasonable request to the authors

## AUTHORS CONTRIBUTION

LVW and EJH designed and supervised the study. BG-G, MLP, RJA, DPC, LJD, AS, OCL and TH-B performed the analyses. CR, PC, FM, CleanUP-IPF Investigators of the Pulmonary Trials Cooperative, HLB, WAF, IPH, SPH, MRH, NH, RBH, RJM, ABM, VN, EO, HP, GS, IS, MDT, MKBW, AA, NK, S-FM, MES, YZ, TEF, MM-M, MN, XRS, JMO, TMM, PLM, CF, IN, DAS, BLY, RGJ and LVW participated in data collection. BG-G, RJA, LVW and EJH wrote the first draft of the manuscript. All authors revised and approved the final version of the manuscript.

