## Supplement for "Association study of human leukocyte antigen (HLA) variants and idiopathic pulmonary fibrosis"

### Supplemental material

Beatriz Guillen-Guio*, Megan L. Paynton*, Richard J. Allen, Daniel P.W. Chin, Lauren J. Donoghue, Amy Stockwell, Olivia C. Leavy, Tamara Hernandez-Beeftink, Carl Reynolds, Paul Cullinan, Fernando Martinez, CleanUP-IPF Investigators of the Pulmonary Trials Cooperative, Helen L. Booth, William A. Fahy, Ian P. Hall, Simon P. Hart, Mike R. Hill, Nik Hirani, Richard B. Hubbard, Robin J. McAnulty, Ann B. Millar, Vidya Navaratnam, Eunice Oballa, Helen Parfrey, Gauri Saini, Ian Sayers, Martin D. Tobin, Moira K. B. Whyte, Ayodeji Adegunsoye, Naftali Kaminski, Shwu-Fan Ma, Mary E. Strek, Yingze Zhang, Tasha E. Fingerlin, Maria Molina-Molina, Margaret Neighbors, X. Rebecca Sheng, Justin M. Oldham, Toby M. Maher, Philip L. Molyneaux, Carlos Flores, Imre Noth, David A. Schwartz, Brian L. Yaspan, R. Gisli Jenkins, Louise V. Wain[#], Edward J. Hollox[#].

*Equal contribution as first authors
[#]Equal contribution as senior authors

### METHODS

#### Study Cohorts

We analysed genomic data from seven previously described independent case-control studies of idiopathic pulmonary fibrosis (IPF). In all seven studies, IPF cases were diagnosed according to the American Thoracic Society and European Respiratory Society guidelines (1,2).

The Study of Clinical Efficacy of Antimicrobial Therapy Strategy Using Pragmatic Design in Idiopathic Pulmonary Fibrosis - University of California Davis (CleanUP-UCD) comprised a total of 469 IPF cases from a randomized clinical trial conducted across 35 US locations and from the University of California Davis (3,4), as well as 2,455 population controls selected from UK Biobank. Both cases and controls were genotyped using the Affymetrix UK Biobank array.

The Colorado study (5) included 1,515 patients with fibrotic idiopathic interstitial pneumonias (fIIP) from US cohorts (National Jewish Health IIP population, InterMune IPF trials, UCSF, Vanderbilt University IIP population, and the National Heart Lung and Blood Institute Lung Tissue Research Consortium) and 2,455 population controls selected so they were genetically similar to the cases. All individuals were genotyped using the Illumina Human 660W Quad BeadChip array.

The Genentech study (6,7) consisted of 813 cases from three IPF clinical trials (ASCEND, CAPACITY, and RIFF) and 3,949 controls from non-IPF clinical trials for age-related macular degeneration, diabetic macular oedema, multiple sclerosis, asthma, and inflammatory bowel disease. Genotypes were obtained from whole-genome sequencing using HiSeq X Ten platform (Illumina Inc.) to an average read depth of 30X.

The Idiopathic Pulmonary Fibrosis Job Exposures Study (IPF-JES) (8) included 416 men from England, Scotland or Wales diagnosed with IPF and 2,465 matching controls (all men) selected from both the IPF-JES study (individuals with an outpatient clinic appointment during the study period) and UK Biobank. Genotyping was performed with the Affymetrix UK Biobank array.

The UK study (9) comprised a total of 612 IPF cases from 9 different centres across the UK and 3,366 matching controls selected from UK Biobank. Cases were genotyped with the Affymetrix UK BILEVE array and controls with the UK Biobank array.

The US study (10) included 541 IPF cases from the University of Chicago, University of Pittsburgh and COMET study, and 542 population controls from the database of genotypes and phenotypes (dbGaP) and the University of Pittsburgh, all genotyped with the Affymetrix Genome-Wide Human SNP 6.0 array.

The UUS study [United States, United Kingdom, and Spain] (11) consisted of 793 IPF cases from 7 study cohorts (ACE, PANTHER, UCD, Chicago, UCSF, PROFILE, and Spain) and 9,999 population controls selected from UK Biobank matching ancestry, sex and smoking distribution. Cases were genotyped with the Affymetrix UK Biobank and Spain Biobank arrays, and controls with the UK Biobank array.

All studies were performed in accordance with The Code of Ethics of the World Medical Association (Declaration of Helsinki) and approved by the appropriate institutional review or Research Ethics Committee.

### SUPPLEMENTARY TABLES

#### Table S1: Sample size of the study

| | Total | Cases | Controls |
|---|---|---|---|
| CleanUP-UCD | 2,924 | 469 | 2,455 |
| Colorado | 6,198 | 1,515 | 4,683 |
| Genentech | 4,762 | 813 | 3,949 |
| IPF-JES | 2,881 | 416 | 2,465 |
| UK | 3,978 | 612 | 3,366 |
| US | 1,083 | 541 | 542 |
| UUS | 10,792 | 793 | 9,999 |
| **Total** | **32,618** | **5,159** | **27,459** |

The number of individuals shown is the result after quality controls.

#### Table S2: Effect allele frequencies in each study

| Variant ID | Position (b38) | NEA/EA | EAfreq Meta | EAfreq / Imp.Q | | | | | | |
|---|---|---|---|---|---|---|---|---|---|---|
| | | | | CleanUP-UCD | Colorado | Genentech | IPF-JES | UK | US | UUS |
| rs4947344 | chr6:32710069 | C/T | 0.286 | 0.293 / 0.991 | 0.290 / 0.987 | 0.281 / NA | 0.281 / 0.991 | 0.283 / 0.997 | 0.290 / 0.984 | 0.280 / 0.992 |
| rs9266618 | chr6:31378389 | A/C | 0.093 | 0.100 / 0.979 | 0.083 / 0.959 | 0.083 / NA | 0.097 / 0.979 | 0.103 / 0.983 | 0.084 / 0.898 | 0.102 / 0.972 |
| rs116450899 | chr6:28764432 | G/A | 0.089 | 0.092 / 0.999 | 0.093 / 0.998 | 0.083 / NA | 0.088 / 0.999 | 0.091 / 0.999 | 0.090 / 0.996 | 0.087 / 0.999 |
| rs138188178 | chr6:30632465 | G/T | 0.043 | 0.044 / 0.989 | 0.044 / 0.99 | 0.046 / NA | 0.04 / 0.985 | 0.043 / 0.995 | 0.036 / 0.894 | 0.043 / 0.987 |

NEA, non-effect allele; EA, effect allele; EAfreq, frequency of the effect allele. Imp.Q, Imputation quality.

### Table S3: Sensitivity analyses of sentinel variants excluding the Colorado study

| Variant ID | Position (b38) | NEA/EA | Results excluding Colorado | | |
|---|---|---|---|---|---|
| | | | OR [95%CI] | P | PPR |
| rs4947344 | chr6:32710069 | C/T | 1.13 [1.07,1.20] | $2.43 \times 10^{-5}$ | 1.24% |
| rs9266618 | chr6:31378389 | A/C | 1.17 [1.07,1.28] | $3.52 \times 10^{-4}$ | 0.63% |
| rs116450899 | chr6:28764432 | G/A | 1.18 [1.08,1.29] | $2.83 \times 10^{-4}$ | 1.88% |
| rs138188178 | chr6:30632465 | G/T | 1.23 [1.08,1.39] | $1.10 \times 10^{-3}$ | 0.88% |

NEA, non-effect allele; EA, effect allele. Associations reaching the Bonferroni threshold ($p$=$4.50 \times 10^{-4}$) are shaded in grey. PPR, posterior probability of replication (obtained with MAMBA, Meta-Analysis Model-Based Assessment of Replicability).

### Table S4: Validation of the *HLA-DQB1*06:02* association with IPF

| Variant ID | Position (b38) | Meta-analysis | | | Excluding Colorado | | |
|---|---|---|---|---|---|---|---|
| | | OR [95%CI] | P | PPR | OR [95%CI] | P | PPR |
| *HLA-DQB1**06:02 | chr6:32663284 | 1.16 [1.09, 1.23] | $5.66 \times 10^{-6}$ | 0.64% | 1.08 [1.00, 1.16] | 0.043 | 0.09% |

PPR, posterior probability of replication (obtained with MAMBA, Meta-Analysis Model-Based Assessment of Replicability).

### SUPPLEMENTARY FIGURES

#### Figure S1. Conditional analysis on rs4947344

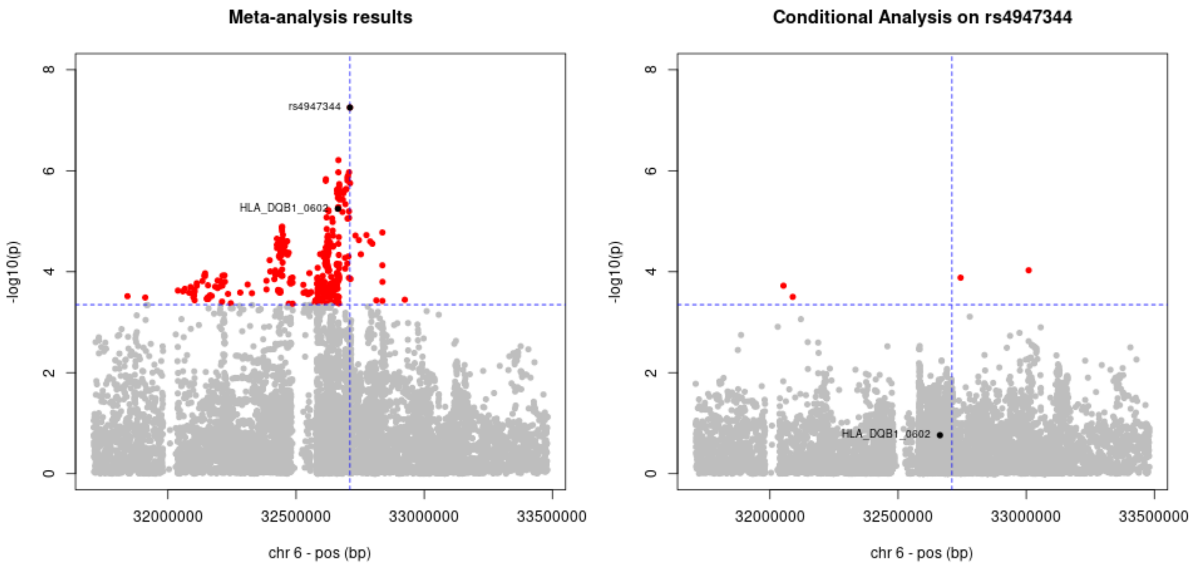


#### Figure S2. Forest plot of the validation analysis results for *HLA-DQB1*06:02*

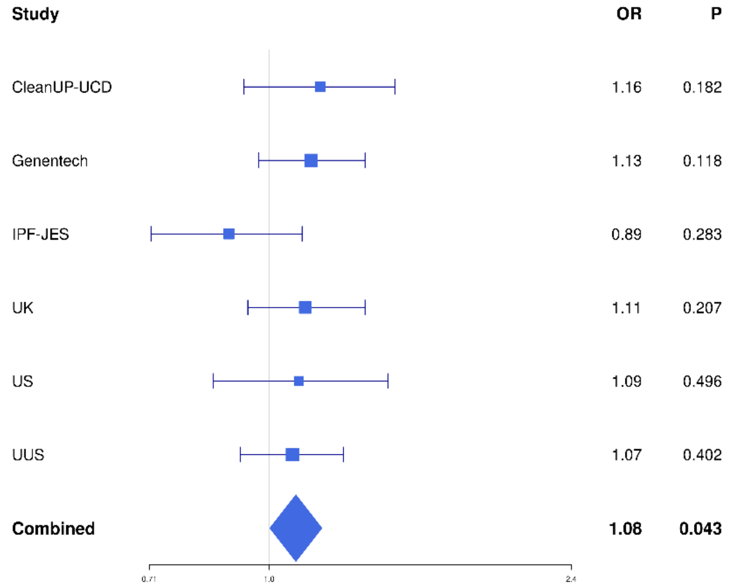
